# Data collection and analysis toolkit for maternal and neonatal health research and surveillance during epidemic threats: a pilot testing and mixed-methods evaluation

**DOI:** 10.64898/2026.10.20.26366068

**Authors:** Veronica Pingray, Karen Klein, Juan Pedro Alonso, Teesta Dey, Luz Gibbons, Alvaro Ciganda, Jose Guilherme Cecatti, Maria Laura Costa, Carla Silveira, Renato T Souza, Jackeline Alger, Jorge García-Aguilar, Pierre Buekens, Deepthy S Melepurakkal, Shivaprasad S. Goudar, Avinash Kavi, Chandrashekhar Kajagar, Manjunath S. Somannavar, Leandre Murhula Masirika, Pacifique Ndishimye, Jean Claude Udahemuka, Brigitte Umutoni, Risa Miyazaki Sankar, Aris Papageorghiou, Adele Winsey, Maria Belizan, Mabel Berrueta

## Abstract

Standardized outcome reporting is essential to generate timely and comparable evidence during epidemics. The World Health Organization (WHO) developed the *Maternal and Neonatal Health during Epidemic Threats Core Outcome Set (MNH-EPI-COS)* and a complementary *Data Collection and Analysis Toolkit* to support its implementation. Before global dissemination, the Toolkit required pilot testing to assess its usability, acceptability, appropriateness, and feasibility across diverse research settings, identify challenges encountered by end users, and inform required improvements and refinements to the Toolkit.

A four-phased mixed-methods evaluation was conducted with five research teams from Brazil, Honduras, India, Rwanda, and the United Kingdom. Teams received training and piloted all Toolkit components using retrospective datasets from previous epidemic contexts. Evaluation methods included structured feedback forms, observation of check-in meetings, and a virtual consultation. Findings informed an iterative process of refinement of the Toolkit components. Data were analysed using descriptive statistics and thematic analysis.

Twenty participants with expertise in epidemiology, clinical research, REDCap administration, and data and management analysis completed the pilot. The REDCap project, R scripts, and supporting materials (reference guide, virtual training session, and videos) were consistently rated as clear, well-structured, and feasible to use. Paper-based case report forms (CRFs) were comprehensive but required refinement, particularly regarding redundancy, and alignment with electronic forms. Teams emphasized the need for multilingual materials, specialized training in R, and strengthened data quality controls to enhance the quality of the evidence generated.

The *WHO MNH-EPI-COS Data Collection and Analysis Toolkit* demonstrated strong usability, acceptability, appropriateness, and feasibility across diverse settings. The findings informed targeted refinements and support the Toolkit’s readiness for broader implementation to strengthen maternal and neonatal health research and surveillance during epidemics. Future evaluations should include prospective, real-time data collection during active outbreaks to further assess and strengthen the Toolkit’s performance under operational epidemic conditions.

## INTRODUCTION

Infectious disease outbreaks and epidemics affect maternal and neonatal health both directly, through disease transmission, and indirectly, through disruption of health services that increase morbidity and mortality. (1–7) Despite these risks, the lack of consistent outcome reporting has long been a major challenge in this context. Heterogeneity in outcome definitions and measurements limits comparability across studies, slows evidence synthesis, and ultimately delays timely and informed public health, clinical, and patient decision-making. (8–10)

To address this gap, and as part of the World Health Organization (WHO) initiative to improve the health and well-being of pregnant women and their babies following infectious disease outbreaks, WHO developed the *Maternal Newborn Health during Epidemic threats Core Outcome Set (MNH-EPI-COS)* through a systematic review, an international consensus process, and a validation survey. (7,11,12) The MNH-EPI-COS defines a minimum set of maternal and neonatal outcomes that should be measured and reported in all studies conducted in the context of ongoing and emerging epidemic threats. It includes seven main maternal outcomes, 11 main neonatal outcomes, seven complementary maternal outcomes, and four complementary neonatal outcomes (**Fig 1**). Main outcomes should be reported as a minimum, alongside any complementary and additional outcomes of interest. Complementary outcomes might not apply to all studies and are important to measure in specific types of studies, settings, or disease outbreaks.

**Figure 1.**
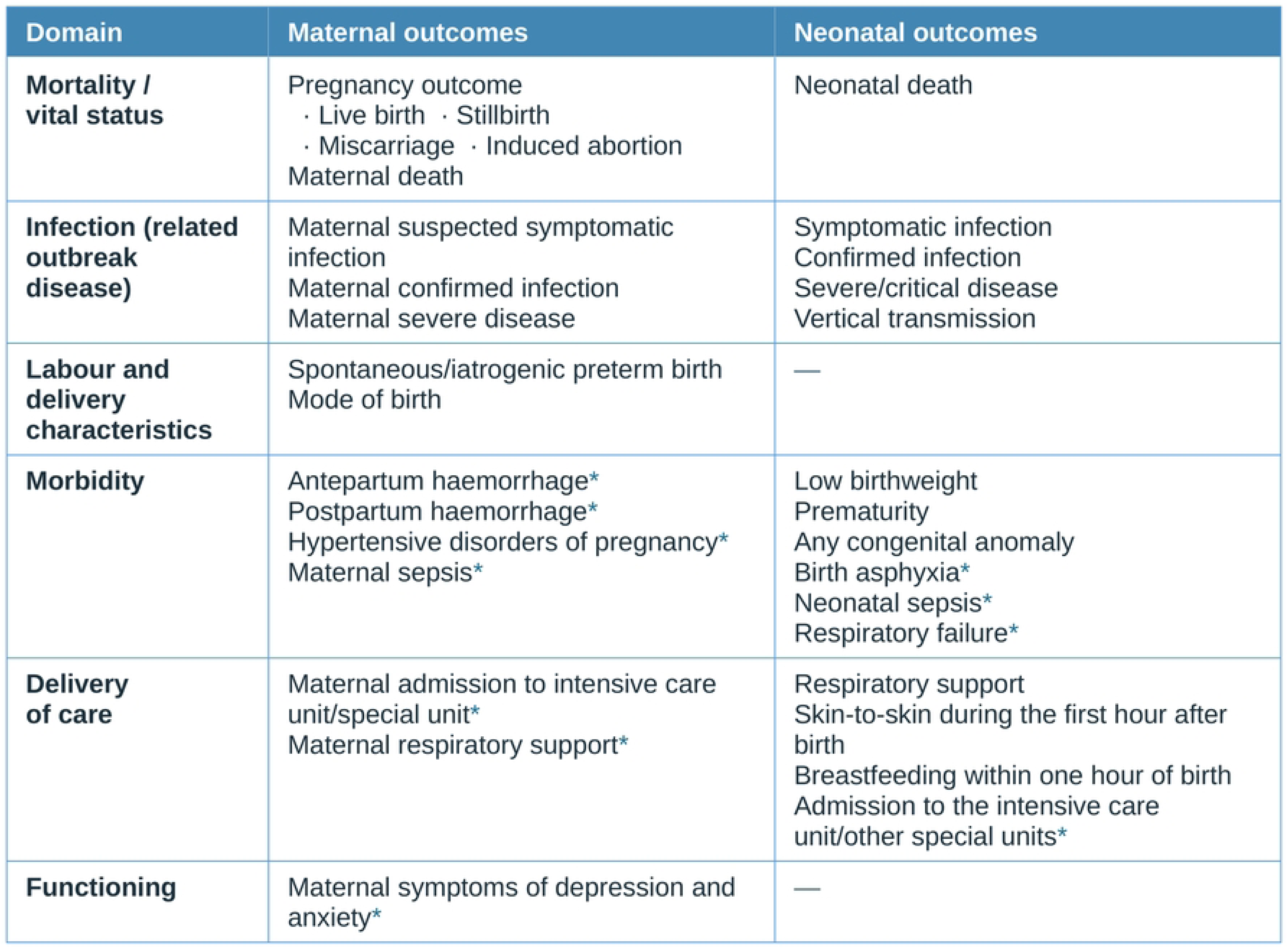
MNH-EPI-COS main and complementary outcomes. Considered as complementary outcomes and proposed to complement the core outcome set when relevant for the setting (depending on resource availability), type of disease and type of research study (e.g., epidemiological, product development, or post-authorization surveillance). (11)

However, developing a core outcome set (COS) alone is insufficient to ensure uptake. Although there are highly successful COS with widespread adoption across their fields, adoption is often slowed by limited awareness, uncertainty about how to use COS, challenges and costs associated with outcome measurement, and researchers’ preference for using their own prioritized outcomes. (13–17) An international online survey conducted in 2024 explored anticipated barriers to MNH-EPI-COS adoption specifically, (18) identifying low awareness of COS in general, perceived high resource requirements, limited understanding of the MNH-EPI-COS, and a lack of clear guidance, practical resources, and validated tools for data collection and outcome measurement as the most frequently cited challenges. (18)

To support MNH-EPI-COS uptake, WHO developed the *MNH-EPI-COS Data Collection and Analysis Toolkit*: a comprehensive set of practical resources for standardized collection, management, analysis, and reporting of maternal and neonatal outcomes in epidemic research and surveillance (**Fig 2**). It is intended to promote the integration of the MNH-EPI-COS across diverse contexts to facilitate the timely and accurate recording and reporting of core outcomes. This Toolkit is designed to reduce the burden on researchers, epidemiologists, and health workers while enhancing the quality, consistency, and comparability of data collected during epidemics. It was developed by experienced technical experts in maternal and neonatal research, epidemiology, REDCap programming, data management, and biostatistics.

**Figure 2.**
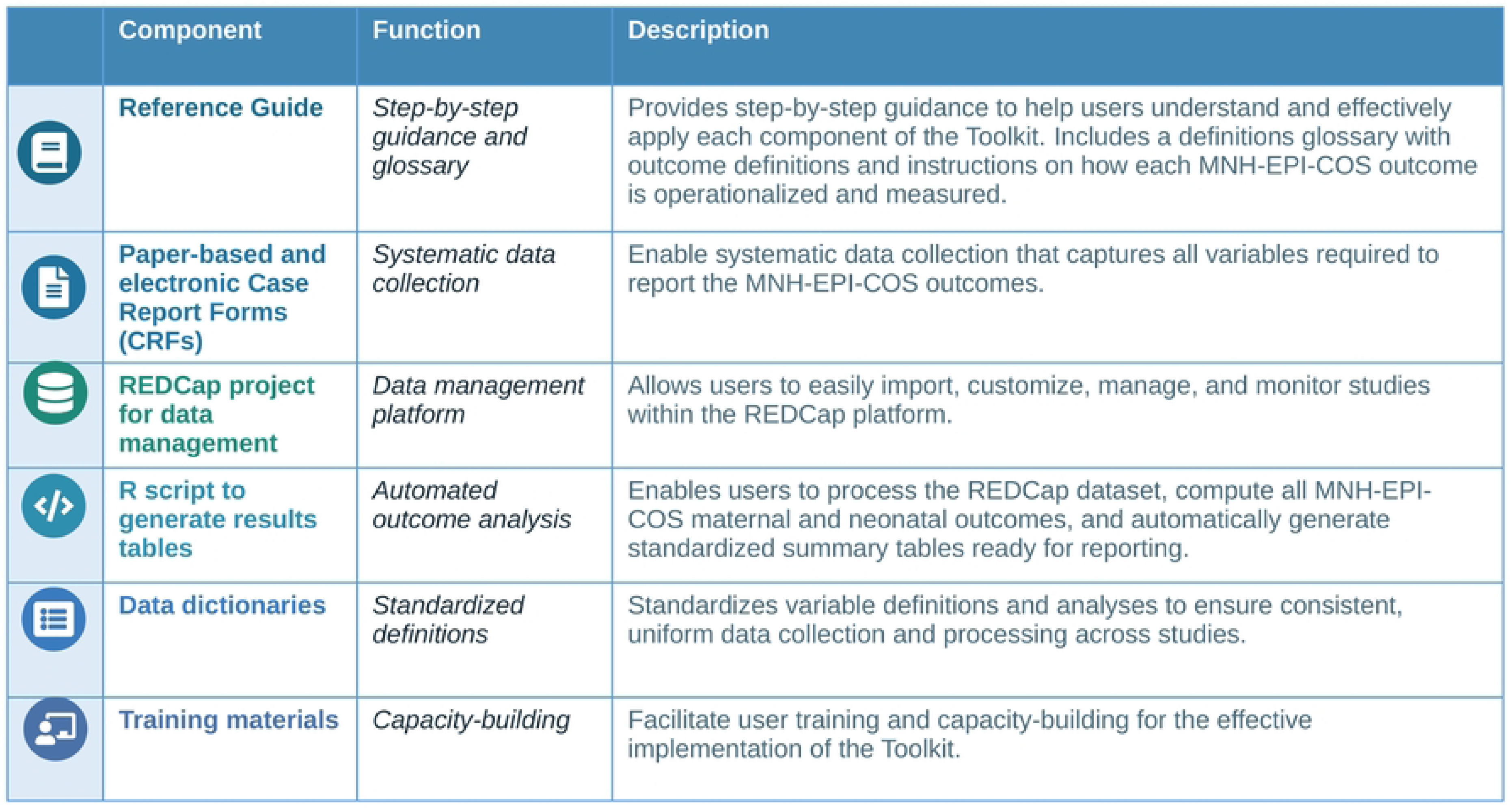
Components of the MNH-EPI-COS Data Collection and Analysis Toolkit.

It is essential to ensure that the *MNH-EPI-COS Data Collection and Analysis Toolkit* performs its intended technical functions, meets the practical needs of its users, and directly supports the standardized collection and reporting of the MNH-EPI-COS. By guiding users to capture core outcomes consistently, the Toolkit is expected to enhance the comprehensiveness, veracity, and accuracy of reported data, thereby strengthening the quality of the outcomes used for evidence synthesis. To achieve this, the Toolkit must enable users to complete data-related tasks effectively, efficiently, and with a high degree of satisfaction, while accounting for the specific contextual factors in which it will be applied. (19) Furthermore, the Toolkit must be perceived as acceptable and feasible across diverse implementation settings, including resource-constrained environments, to ensure reliable and comparable outcome reporting across studies and surveillance efforts. The aim of this project was to pilot-test the *MNH-EPI-COS Data Collection and Analysis Toolkit* and evaluate its usability, appropriateness, acceptability, and feasibility among research and epidemiological teams. In addition, the project sought to identify users’ reported challenges encountered during Toolkit implementation, define areas requiring improvements, and incorporate them into the final version of the Toolkit.

## METHODS

We conducted a mixed-methods evaluation to pilot-test the *MNH-EPI-COS Data Collection and Analysis Toolkit*, with the participation of research and epidemiological teams (i.e., researchers, epidemiologists, data managers, data analysts, and REDCap administrators). The evaluation was implemented in four sequential phases: (1) preparation and training; (2) pilot implementation of all Toolkit components using datasets generated during previous outbreaks, epidemics, and pandemics; (3) evaluation and feedback; and (4) Toolkit refinement and finalization (**Fig 3**).

**Figure 3.**
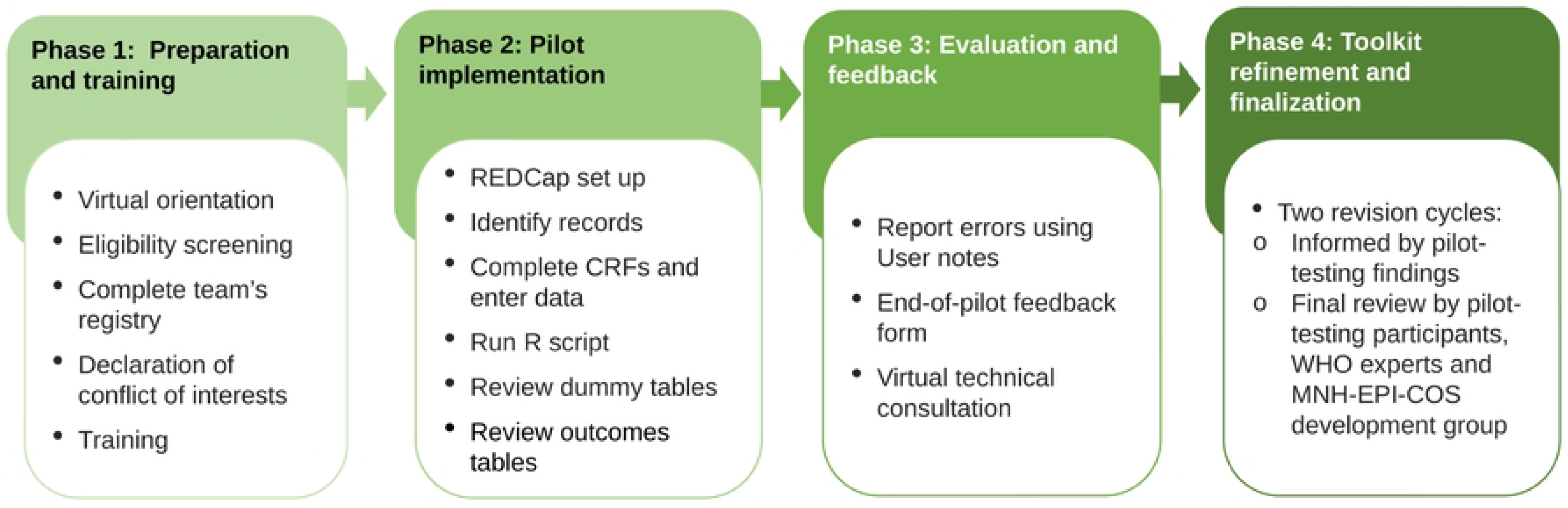
Project phases and main activities.

### Settings and participants

Sixteen potential collaborator groups with a history of conducting maternal and neonatal health research or surveillance during infectious disease outbreaks were identified through prior partnerships with the coordination team. Seven teams accepted to participate in an orientation session, which provided an overview of the pilot and evaluation project. Following the session, between September 10 and 18, 2025, interested teams completed a structured form (**S1 File)** to verify eligibility and to provide descriptive information about their teams and datasets. Eligibility criteria included: (a) access to retrospective datasets containing maternal, neonatal, and infectious disease data from studies conducted during outbreaks, epidemics or pandemics; (b) availability of variables sufficient to report at least 50% of MNH-EPI-COS main outcomes, (c) inclusion of at least one of the following populations: pregnant women at any trimester of gestation, women in labour or post-pregnancy, and neonates of any age between 0 and 28 days after birth; and (d) data of a minimum of 10 participants with suspected or confirmed WHO prioritised infection with epidemic potential. (20) In addition, teams were eligible if they demonstrated prior experience in maternal and neonatal health research or epidemiological surveillance during outbreaks, sufficient technical capacity to use REDCap and run R, and obtained relevant institutional approvals.

A convenience sample of five research teams from Brazil, Honduras, India, Rwanda, and the United Kingdom, that met the eligibility criteria were included, while ensuring representation across diverse geographical regions and infectious diseases under study. These teams included multidisciplinary experts in epidemiology, clinical research, REDCap, data management, and data analysis, reflecting the intended end users of the Toolkit.

### Outcomes

We evaluated the usability, acceptability, appropriateness, and feasibility of both the individual components of the *MNH-EPI-COS Data Collection and Analysis Toolkit* and its overall performance. This evaluation included identifying benefits and challenges and collecting user recommendations for improvement. These outcomes were selected because the Toolkit is intended for rapid uptake during epidemics, where tools must be not only technically robust but also intuitive, practical, and contextually appropriate. Together, these constructs represent key early-stage implementation outcomes commonly used to determine whether an intervention is suitable for broader rollout. (21)

Usability was assessed using items adapted from the System Usability Scale (SUS), focusing on task completion, instruction clarity, and ease of use. (22) Acceptability, appropriateness, and feasibility were measured using items adapted from the Implementation Outcome Measures (AIM, IAM, and FIM) developed by Weiner and colleagues, building on Proctor et al. framework (21,23), tailored to the pilot context while remaining aligned with the original instruments.

Experts also documented any technical errors or limitations encountered during use, enabling a comprehensive assessment of the Toolkit practical performance and perceived suitability that could directly inform revisions (**Table 1**).

**Table 1.** Evaluation dimensions and items used in the pilot-testing evaluation.

| Outcome Dimension | Item (customizable by Toolkit component) |
| --- | --- |
| Usability – Task completion | Our team completed the (component). |
| Usability – Clarity | The instructions for completing/using the (component) were clear and concise. |
| Usability – Ease of use | The (component) was easy to use in practice. |
| Usability – Satisfaction | Our team is satisfied with the (component). |
| Acceptability | Our team would recommend using this (component) in relevant research settings. |
| Appropriateness | The (component) seems suitable for its intended purpose. |
| Feasibility | The (component) seems feasible to implement in research studies. |
| Open-ended feedback | Please describe the main positive aspects / negative aspects or challenges of the (component), based on your team's experience. |

### Main activities

#### Phase 1. Preparation and training

The process began with a virtual orientation session to introduce the objectives of the pilot, the background and rationale of the MNH-EPI-COS and the Toolkit, and the scope and expected outputs of the evaluation. Teams then completed a structured form capturing team composition, expertise, dataset characteristics, and available infrastructure, used to verify eligibility, tailor implementation scenarios, and anticipate support needs.

All five teams subsequently took part in a 2.5-hour training session establishing a common baseline understanding of the Toolkit and covering the structure and use of all components— reference guide, CRFs, REDCap project, and R scripts—including guidance on data extraction, CRF completion (paper-based and electronic), REDCap configuration, and R script execution. The session was delivered by the coordination team and the technical experts who developed the Toolkit, recorded, and made available for teams to revisit throughout the pilot.

#### Phase 2. Pilot implementation of the Toolkit

In Phase 2, teams applied the Toolkit to their own retrospective datasets, following the reference guide and training materials. They created a REDCap project on institutional servers using the standardized template, customized selected variables (e.g., infection type, clinical signs and symptoms) to their dataset, and identified at least 10 eligible records of suspected or confirmed infection and, where possible, an equivalent set of controls. Tailored testing scenarios—reflecting maternal infection during pregnancy, neonatal infection, and pregnancy outcomes such as miscarriage, abortion, stillbirth, or multiple births—were assigned to capture a wide range. Teams completed the CRFs using both paper-based and electronic formats. After data entry, teams exported their datasets and ran the R scripts to generate standardized output tables (**Fig 4**).

**Figure 4.**
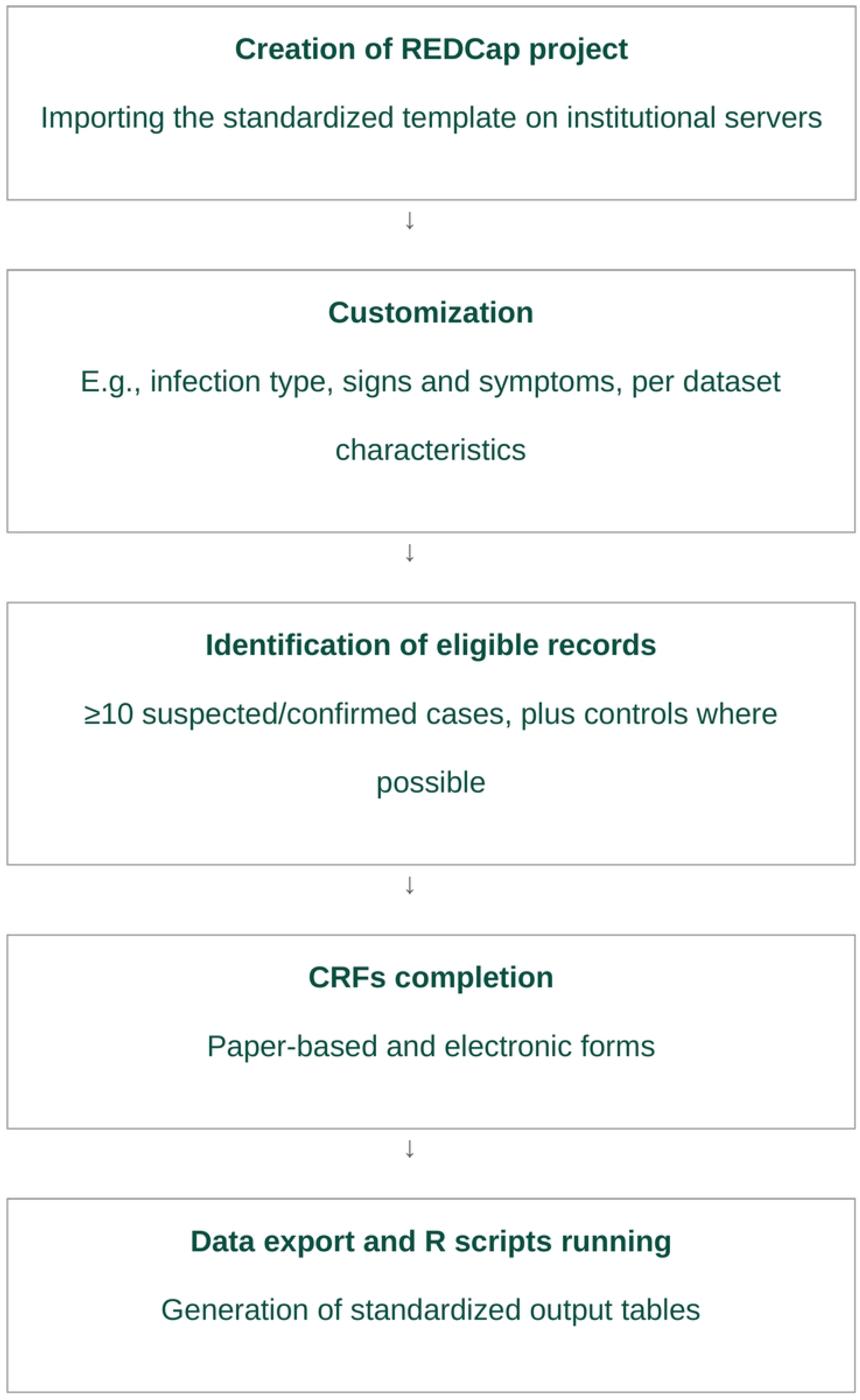
Phase 2 workflow.

Throughout, participants documented task completion, errors, difficulties, time requirements, and usability/workflow observations using structured User notes (**S2 File)**. Regular check-in meetings provided technical support and captured additional implementation challenges.

#### Phase 3. Evaluation and feedback

After completing pilot activities, teams collectively completed a piloted, structured end-of-pilot feedback form (**S3 File**) combining a 5-point Likert scale (“Strongly disagree” to “Strongly agree”) with open-ended questions on usability, acceptability, appropriateness, feasibility, and areas for improvement; each team submitted a single consolidated response incorporating input from members with diverse expertise.

All teams also took part in a 2-hour, semi-structured virtual consultation facilitated by an experienced social scientist (**S4 File**), aimed at reviewing pilot feedback to identify priority Toolkit improvements and exploring additional strategies to strengthen maternal and neonatal research and surveillance during future epidemics.

#### Phase 4. Toolkit refinement and finalization

The final phase synthesized all feedback to inform Toolkit refinement, drawing on two sources: (a) pilot-testing findings from participating research and surveillance teams, and (b) feedback from WHO experts and from experts who had contributed to MNH-EPI-COS development. (11)

Expert review to validate the content was undertaken both internally at WHO and with external experts who had participated in the development of the MNH-EPI-COS Toolkit. This involved two groups: seven WHO experts spanning biostatistics, research capacity strengthening, clinical trials and pharmacovigilance, epidemic response, infectious disease, immunization, reproductive health, maternal care, and paediatric care, who provided written feedback and discussed it in a virtual meeting; and seven international experts from the MNH-EPI-COS development group—covering maternal, perinatal, and neonatal health, infectious disease, public health and emergency response, and civil society—who reviewed the post-pilot Toolkit version and provided written feedback. This revised version was also shared with pilot participants to assess whether the changes had adequately addressed the challenges they had identified.

All feedback was reviewed and classified by the coordinating team. Two coordinating team members coded and grouped all suggested improvements by issue type; these were then discussed with the full coordinating team and technical experts and jointly classified as either “proceed with the change” (addressed in the current revision cycle) or “out of scope” (deferred to a subsequent phase). Suggestions classified as “proceed with the change” were incorporated into the final version of the Toolkit.

### Data analysis

Quantitative data from the end-of-pilot form were summarized using descriptive statistics and, where appropriate, graphically represented using heatmaps. Categorical variables were described using absolute and relative frequencies.

Qualitative data from open-ended responses were analysed using thematic analysis to identify prominent themes, patterns, and key messages related to user experience and implementation. In addition, notes from observation of the check-in meetings and user feedback (user notes) were systematically compiled and analysed to capture detailed insights into implementation processes and challenges. Transcripts and field notes from the virtual consultation were also analysed thematically to explore further cross-cutting challenges and recommendations for improvement across diverse settings.

Quantitative and qualitative findings were triangulated and are presented in an integrated manner for each Toolkit component and for the Toolkit as a whole, with open-ended responses used to elaborate and contextualise quantitative findings where relevant.

Quantitative data were analyzed using Stata version 14.0 (StataCorp, College Station, TX, USA), and qualitative data using ATLAS.ti.

## ETHICAL CONSIDERATIONS

This project assessed the usability of a Toolkit components by collecting expert feedback from research and surveillance teams with relevant technical expertise. The evaluation relied exclusively on secondary, de-identified, retrospective data from pre-existing databases. All original studies had undergone IRB review and approval. Participating teams independently conducted internal analyses to evaluate the Toolkit components, without transferring original study data to the coordination team. Consequently, formal ethics approval was generally not required. However, the Rwanda and India teams submitted the protocol and annexes to their local IRBs and were exempted from full review.

The pilot adhered to recognized standards of good clinical and research practice (GCP), ensuring the protection of all individuals involved and the integrity of the data. Before contributing, participants completed the standard WHO declaration of interest form, disclosed any potential academic, professional, financial, or other conflicts of interest, and provided electronic consent.

## RESULTS

### Participants and datasets characteristics

Five teams with 20 experts from Brazil, Honduras, India, Rwanda, and the United Kingdom participated in this evaluation. Most participants were researchers and data managers, while two participants identified as IT/REDCap administrators and biostatisticians/data analysts. Most had more than 10 years of experience (**Table 2**).

**Table 2.**
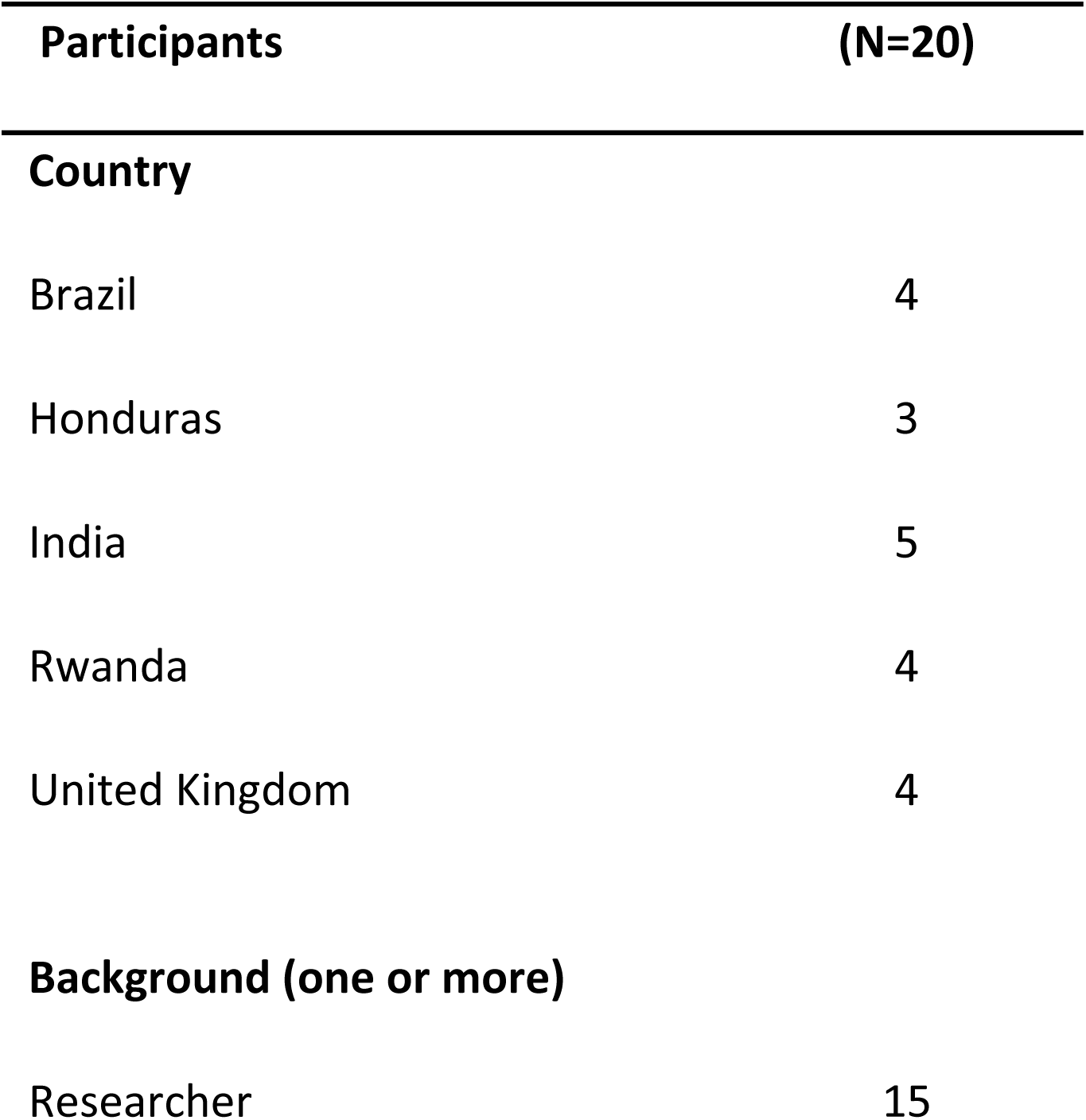
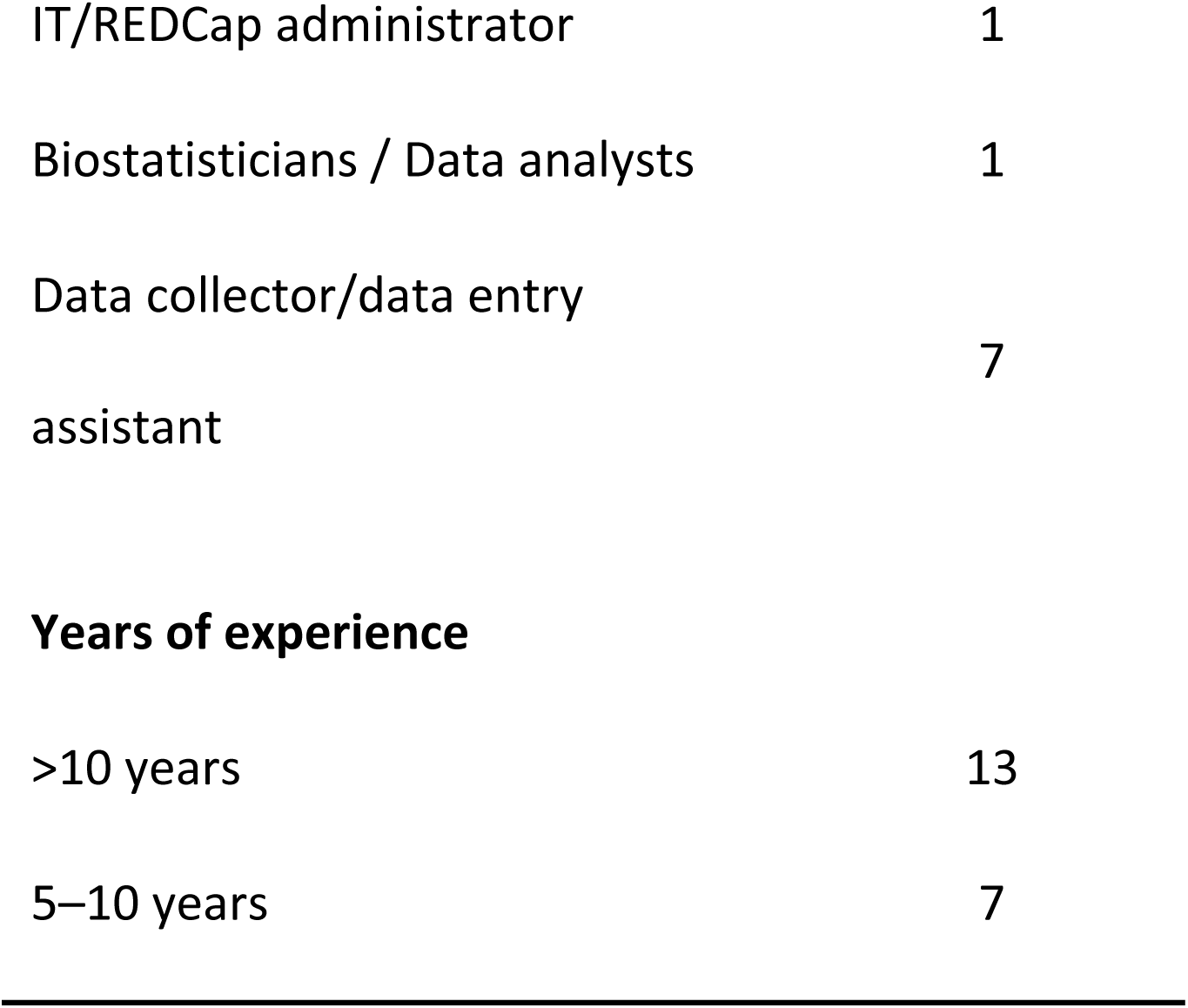
Evaluation of participants’ characteristics.

| Participants | (N=20) |
| --- | --- |
| **Country** |  |
| Brazil | 4 |
| Honduras | 3 |
| India | 5 |
| Rwanda | 4 |
| United Kingdom | 4 |
| **Background (one or more)** |  |
| Researcher | 15 |
| IT/REDCap administrator | 1 |
| Biostatisticians / Data analysts | 1 |
| Data collector/data entry assistant | 7 |
| Years of experience |  |
| >10 years | 13 |
| 5–10 years | 7 |

Teams worked with datasets collected during COVID-19, H1N1 influenza, Zika, and Mpox epidemics. One of the participating teams conducted the pilot using two separate datasets collected during two different pandemics (COVID-19, H1N1 influenza). All studies were observational with a comparison group, and most included both maternal and neonatal outcomes (**Fig 5**). All datasets contained at least 50% of MNH-EPI-COS outcomes, except the Zika dataset, which included 43% of MNH-EPI-COS outcomes, but it was retained due to the relevance of congenital infections (**S1 Tables and S2 Table**).

**Figure 5.**
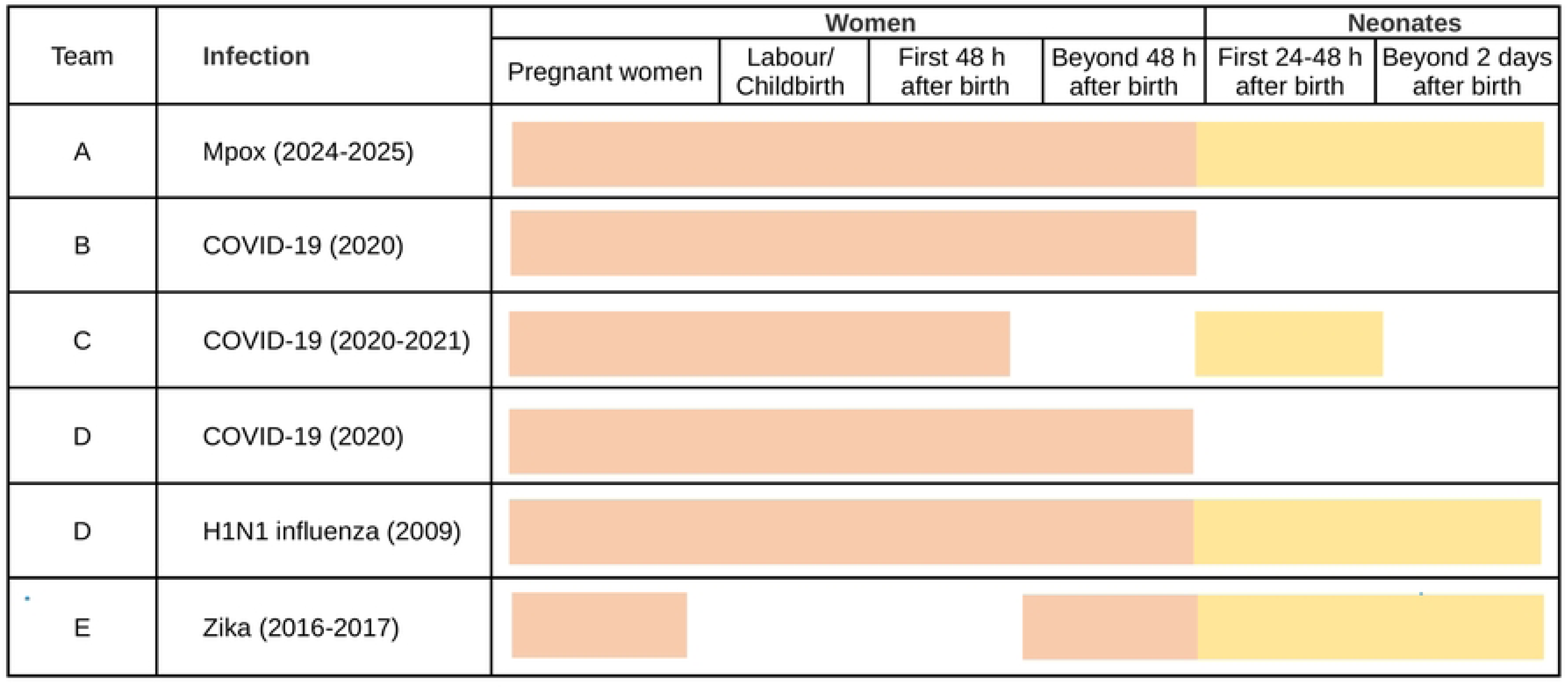
Participating research teams and dataset characteristics.

### Experience using the *MNH-EPI-COS Data collection and analysis Toolkit*

Across all Toolkit components, the evaluation demonstrated consistently positive results among the five participating teams, with only minor variability between teams and components. Most evaluation items were rated highly, indicating that teams were able to complete key tasks, found the instructions clear, and considered the components easy to use and suitable for their intended purposes (**Fig 6**).

**Figure 6.**
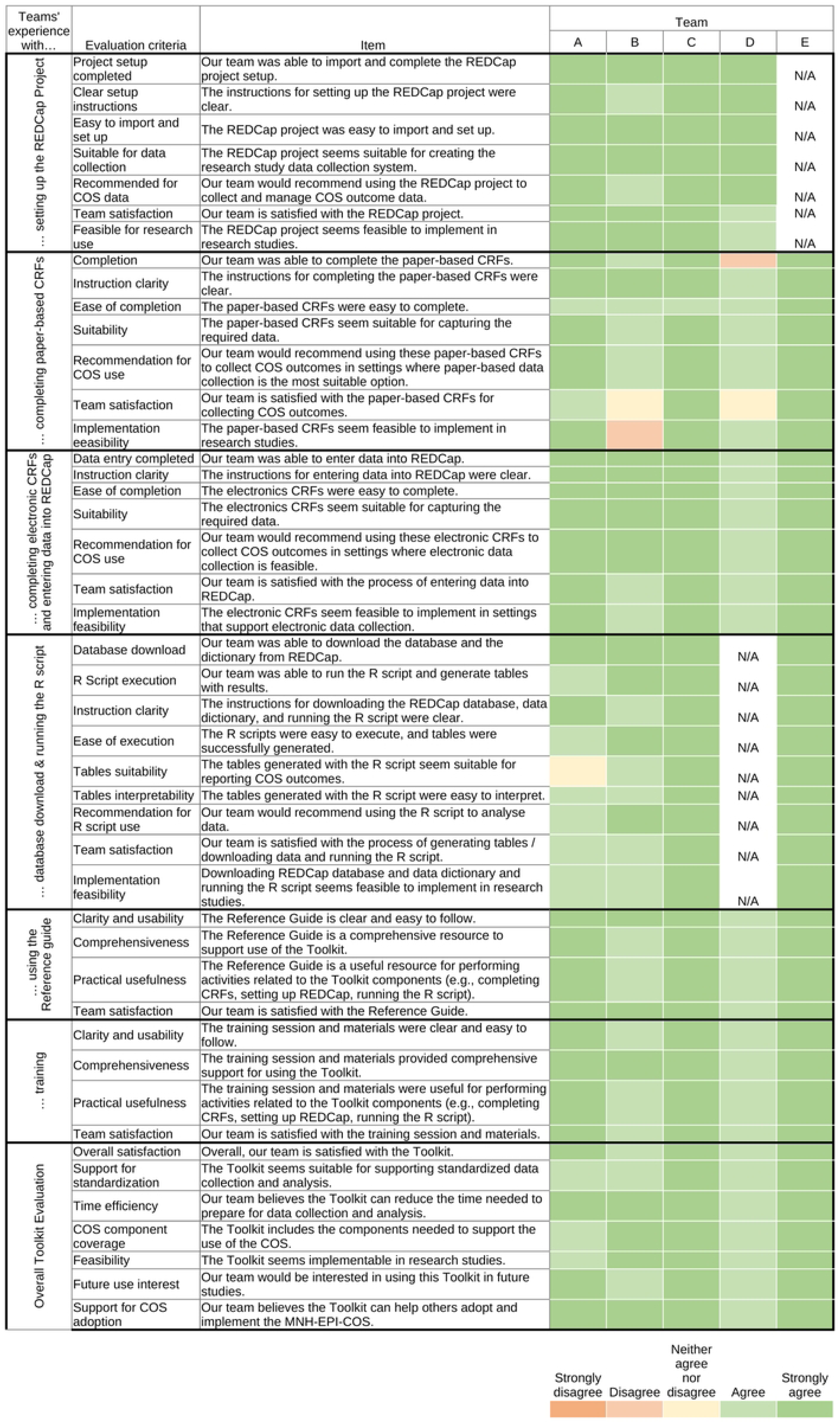
Participating teams’ experiences implementing the *MNH-EPI-COS Data Collection and Analysis Toolkit.* Note: N/A= not applicable.

#### Setting up the REDCap project

To start, teams were asked to follow the instructions in the reference guide to use a REDCap XML template to set up a fully structured REDCap project preloaded with the CRFs. One team did not perform the REDCap project setup due to a lack of an institutional license; in this case, the coordination team set up the project on their behalf, and they did not evaluate this component.

Participating teams consistently reported a positive experience setting up the REDCap project, agreeing that it could be successfully completed, that instructions were clear, and that the system was easy to import and program. In open comments, teams valued the platform’s online accessibility for remote work without relying on physical storage, and its role-based access controls as particularly relevant for data security and confidentiality during outbreak research. The project was consistently rated as suitable for creating a research data collection system aligned with MNH-EPI-COS. Teams also noted its flexibility and compatibility with existing datasets, and versatility across different diseases. Teams reported high satisfaction and would recommend its use for collecting and managing COS outcomes data. The REDCap project was also considered feasible for implementation in research studies, and consider it suitable across diverse settings, particularly where teams operate under time pressure or with limited infrastructure.

#### Using paper-based Case Report Forms

Paper-based CRFs, completed with data extracted from teams’ own databases, showed greater variability in ratings than other components (**Fig 6**). Most items—clarity of instructions, ease of completion, suitability, and likely recommendation—were rated positively; teams described the forms as well-designed, comprehensive, and logically structured, with standardized definitions supporting consistent completion and print-ready formatting valued for adaptability. However, two of the five teams gave neutral or negative ratings for completion, satisfaction, and feasibility, attributing this to the absence of formatting to highlight skip logic—which left all sub-questions visible regardless of relevance and increased error risk—and to redundancy and ambiguity in some response options. Teams recommended adding visual cues and flowcharts to indicate which CRFs apply to specific scenarios, and emphasized the need for multilingual versions.

#### Completing electronic CRFs and entering data into REDCap

All teams successfully completed data entry and agreed that instructions were clear and that electronic CRFs were easy to complete and suitable for the required data (**Fig 6**). Required fields and branching logic were valued for guiding users and minimizing errors, while automated form display (showing only relevant forms per scenario) and automatic ID management reduced redundancy and prevented record duplication. Teams reported high satisfaction and would recommend electronic CRFs where electronic data collection is feasible, though they noted difficulties reviewing and verifying data within REDCap, minor inconsistencies with the paper-based version, and connectivity barriers in low-resource settings. As with other components, teams called for multilingual versions.

#### Executing the R Script and generating results tables

Teams downloaded their REDCap database and data dictionary and ran the R script to generate results tables; one team could not evaluate this component due to analyst unavailability. The remaining four teams completed the process successfully and agreed that instructions were clear, the script was easy to use, and they would recommend it for data analysis (**Fig 6**), describing the workflow from export to table generation as straightforward and efficient. Generated tables were rated suitable for reporting COS outcomes and easy to interpret; automated generation was noted to reduce manual workload and transcription errors, supporting a more streamlined, reproducible analysis. One team gave a neutral rating on table suitability, citing an incomplete dataset that limited full script testing. Teams reported satisfaction and feasibility overall, though open comments noted that successful execution requires basic R familiarity and would benefit from additional training resources.

#### Using supporting materials

Teams consistently agreed that the Reference Guide and training were clear, comprehensive, and useful for completing Toolkit activities, reporting satisfaction with both (**Fig 6**). Step-by-step instructions and standardized definitions were noted to support consistent implementation with minimal external assistance, though teams suggested adding diagrams, flowcharts, and examples to clarify more complex processes in the Reference Guide. Recorded training sessions were valued for allowing participants to revisit content and accommodate time-zone differences, though some found the REDCap and R sections technically complex when prior experience was limited. As with other components, teams called for translation into additional languages, noting that English-only materials limit usability in many outbreak-affected contexts.

#### Overall Toolkit evaluation

All teams agreed that they were satisfied with the Toolkit, that it supports standardized data collection and analysis while reducing preparation time, that it includes the components needed to report the MNH-EPI-COS, and that it is implementable, worth using in future studies, and helpful for others adopting the MNH-EPI-COS (**Fig 6**). In open comments, teams described it as a practical set of resources that addresses a clear methodological gap in maternal and perinatal health research, bringing together essential components in a ready-to-use format that reduces preparation time and supports consistent reporting across studies. Its flexibility -accommodating different infectious diseases and including both maternal and neonatal outcomes-was considered an important strength, as were the standardized workflows and built-in features that help reduce errors and improve the efficiency of data entry and analysis.

Suggestions for improvement focused on refining the CRFs and expanding language options. During the virtual consultation, teams identified additional components or forms of support should be incorporated into the Toolkit. Participants underscored the importance of timely evidence synthesis to inform decision-making in the context of public health emergencies, as well as the need for systematic data quality procedures during study implementation. Drawing on experiences analyzing health data during the COVID-19 pandemic, participants reported inconsistencies in data collection across settings and the absence of systematic quality assurance procedures. These gaps were associated with potential biases—such as selective recruitment with wide geographical representation and cultural variability compromised the reliability of the information generated. This underscored the need for standardized protocols and procedures to minimize bias during data collection and strengthen the consistency and reliability of analyses.

### Other implementation strategies to strengthen maternal and neonatal research capacity during epidemics

During the virtual consultation, participants reflected on the strategies outlined in the WHO Roadmap for research on maternal and perinatal health in the context of epidemic threats to identify priorities for strengthening future research and surveillance. (7)

Discussion centered on the need for harmonized research practices, stronger surveillance infrastructure, and coordinated international strategies for effective emergency response. Participants valued sustained international collaborations that maintain health registries and adapt during emergencies—accelerating evidence generation by building on existing infrastructure—and considered strengthening these networks essential for both country-level and multinational analyses. Self-paced online courses were seen as an effective strategy for preparedness and capacity building, supporting consistent implementation of harmonized practices across settings. Given the operational challenges of initiating research once an outbreak begins, participants agreed that preparedness activities—including protocol harmonization and development of ready-to-use tools—must occur before the next public health emergency.

### Improvements addressed and priorities identified for subsequent phases

A range of issues spanning content clarity, consistency, usability, and technical functionality were identified across Toolkit components; most were addressed, while selected areas were deferred to future phases (**S3 Table** details pilot participants’ and experts’ suggestions and their disposition).

Based on pilot participants’ feedback, within the CRFs, redundant or overlapping questions were revised, response options were clarified, and flowcharts were improved to support navigation, along with the addition of a limited number of fields; wording and skip-logic inconsistencies between paper-based and electronic versions were also harmonized. The REDCap module was enhanced with auto-generated infant forms and dedicated quality control rules to enable systematic data quality monitoring and discrepancy management, allowing research teams to identify and resolve missing data, inconsistencies, and data entry errors throughout the data collection process. Minor variable-coding errors in the R scripts were corrected, and guidance on output file locations was clarified. Broader needs for R training and customized analysis guidance were considered beyond the scope of this phase; and connectivity limitations and need for translations were identified as priorities for future phases.

Based on the expert review feedback, we revise the R script to report testing rate and timing, aligned key outcome definitions across components, incorporated validation rules between laboratory results and infection status, and added clarifications on ICD coding. (24) The Reference Guide was also updated to clarify the Toolkit’s scope, improve tables and figures titles, and incorporate GCP/ethics guidance. (25–27)Finally, the training package was revised to clarify prerequisites, duration, and modality. Proposed refinements to outcome definitions and measurements, as well as vaccine-safety-surveillance additions, were disregarded, as they diverged from the agreements reached through the consensus process. However, it was clarified that the variables currently included in the CRFs allow complete reporting of the MNH-EPI-COS main and complementary outcomes, and that researchers and epidemiologists may add new variables if needed to address other research questions. Missing-value recoding was revised, while requests for confidence intervals and further documentation of composite variables were deferred to future development.

Overall, both the pilot participants and the experts feedback were very informative and resulted in multiple revisions to enhance the Toolkit, while also identifying key areas for further development.

## DISCUSSION

### Main findings

Multidisciplinary teams from five countries — representing high-income, upper-middle-income, and lower-middle-income settings — were able to implement all Toolkit components using retrospective datasets spanning four different epidemic contexts (COVID-19, H1N1 influenza, Zika, and Mpox). Ratings were consistently positive across teams and components, and qualitative findings corroborated and elaborated these assessments, collectively indicating that the Toolkit is ready for broader implementation following targeted refinements.

The REDCap project, R scripts, and supporting materials — including the reference guide and training sessions — were the strongest-performing components, rated highly across all evaluation dimensions: task completion, clarity of instructions, ease of use, suitability, acceptability, and feasibility. Electronic CRFs also performed well; features such as branching logic, required fields, and automated ID management were consistently highlighted as practical strengths that reduce data entry errors and improve workflow efficiency. Paper-based CRFs, while described as comprehensive and well-structured, showed greater variability across teams and were the only component where two teams reported neutral or negative ratings, primarily related to satisfaction, and feasibility. Challenges concentrated around the absence of clear skip logic formatting, redundancy in some variables, and the absence of multilingual versions. The Toolkit as a whole was viewed as filling a critical methodological gap by providing a standardized, ready-to-use package for collecting and reporting maternal and neonatal outcomes during epidemics — reducing preparation time and supporting consistent reporting across diverse contexts.

The pilot additionally identified cross-cutting priorities for strengthening the Toolkit, including REDCap-based data quality control procedures, introductory R training resources, and translation into additional languages.

### Interpretation

Evidence from prior COS implementation efforts consistently highlights that even well-designed standardized tools face barriers related to perceived burden, limited awareness, and the absence of practical implementation resources. (14–16) Our findings should be interpreted in light of recent operational evidence from the multicountry WHO COVID-19 pregnancy cohort study, which demonstrated that generating robust maternal and perinatal data across diverse LMIC settings during a pandemic is feasible. (28) The study also describes the challenges of building research infrastructure once an emergency is underway. Its authors therefore concluded that CRFs of core outcomes and generic protocols should be available in advance, together with research sites ready to activate data collection when an outbreak occurs. The Toolkit is designed to ease implementation, improve data quality, and accelerate analysis through automated R scripts. However, how well it works under real-time epidemic conditions remains to be established, reinforcing the need for prospective evaluation during an active outbreak.

The Toolkit was specifically developed to address these barriers, and the pilot results suggest that it largely succeeds in doing so for the components evaluated. The residual challenges identified — particularly multilingual accessibility and R proficiency — are important equity considerations that will need to be systematically addressed to ensure that the Toolkit can be used by the research and surveillance teams in the low-and middle-income countries most affected by epidemic threats.

The finding that electronic CRFs outperformed paper-based CRFs in terms of usability and feasibility is consistent with evidence from other studies evaluating hybrid data collection approaches in research settings, which generally find that electronic systems — when supported by adequate infrastructure — reduce error rates, improve data completeness, and increase user satisfaction. (29–31) The connectivity and institutional-access caveats raised by teams reflect well-known LMIC infrastructure challenges and underscore the priority of developing robust offline solutions for future Toolkit development.

Despite the importance of practical resources for COS uptake, evaluations of such resources are rare in the literature. The field has primarily focused on developing guides and user manuals for specific COS — such as the users’ guide developed by the HOME initiative for the atopic dermatitis COS, which addressed questions about instrument access, training requirements, and appropriate application — but without systematic empirical evaluation of usability or feasibility. (32) A recently published commentary explicitly highlighted this gap, proposing that dissemination and implementation science frameworks should be applied more systematically to support COS uptake and scale. (33) The current study provides a systematic evaluation of a dedicated data collection and analysis Toolkit designed to operationalize a COS, using validated implementation outcome measures.

### Strengths and limitations

This evaluation has several strengths. It employed a structured mixed-methods design that combined quantitative measures with qualitative data from multiple sources — structured feedback forms, observation of check-in meetings, user notes, and a semi-structured virtual consultation — enabling triangulation of findings and a comprehensive understanding of both the magnitude and the nature of the implementation challenges encountered. The geographic and disciplinary diversity of participating teams, spanning five countries across three continents and four epidemic contexts, enhances the contextual breadth of the findings. Finally, the iterative refinement process, in which pilot findings directly informed two structured revision cycles with input from pilot participants, MNH-EPI-COS development group members, and WHO technical experts, ensures that the final Toolkit is evidence-based and end-user-informed.

Several limitations should also be acknowledged. First, and most importantly, the evaluation relied on retrospective datasets from previous epidemics rather than prospective, real-time data collection during an active outbreak. This may not fully reflect the operational pressures that teams would face when implementing the Toolkit during an ongoing public health emergency — the setting for which it was ultimately designed. The performance of the Toolkit under conditions of active epidemic response, including rapid staff onboarding, high patient volumes, and institutional disruption, remains to be established. Second, while participating teams represented diverse geographic regions and epidemic contexts, all met minimum technical requirements — including the ability to use REDCap and run R — and had prior experience in maternal and neonatal health research. This selection criterion, necessary to ensure evaluability of all Toolkit components, means that the findings may not generalize to research or surveillance teams with lower baseline technical capacity. Third, the evaluation was conducted with a small convenience sample of five teams, which, while appropriate for a pilot study, may not capture the full range of contextual variation in implementation experiences. Fourth, some components that were identified as important for broader implementation — multilingual versions, offline REDCap functionality, and a complete R training module — were outside the scope of this pilot and could therefore not be evaluated, leaving a gap in the evidence base for implementation in some settings.

### Implications for practice

These findings support the broader dissemination of the Toolkit as a practical, ready-to-use resource for standardized maternal and neonatal data collection during epidemics. Prospective users—research teams, epidemiologists, and public health surveillance professionals—can be reassured that it has been systematically evaluated and refined across diverse settings and epidemic contexts, with consistent performance on usability, acceptability, appropriateness, and feasibility. Although ready to use, the Toolkit does require initial training; in this pilot, a single 2.5-hour session, recorded for later reference, was sufficient for experienced teams to implement all components.

Teams relying primarily on paper-based data collection should note that the CRFs, while comprehensive and well-structured, require careful navigation, and that the revised reference guide’s flowcharts and skip-logic guidance are essential companions. Teams with limited R experience will benefit from supplementary training before running the analytical scripts, and electronic CRFs are preferable where REDCap connectivity is possible. Because the Toolkit is currently available only in English, non-English-speaking settings will need translated CRFs and supporting resources. Beyond these operational considerations, integration with existing in-country data collection and health information systems should be considered to ensure sustainability.

### Implications for research

The most immediate priority is a prospective evaluation of the Toolkit during real-time data collection in the context of an active epidemic or outbreak. Such an evaluation would assess Toolkit performance under genuine operational conditions, including time pressure, rapid staff onboarding, and the data quality challenges. It would also generate evidence on the Toolkit*’s* downstream impact — specifically, whether its use improves the completeness, consistency, and comparability of reported MNH-EPI-COS outcomes across studies.

Longer-term research on the adoption and sustainability of the Toolkit will ultimately be required to evaluate whether it achieves its intended goal of harmonizing outcome reporting across maternal and neonatal research studies conducted during epidemic threats globally.

## CONCLUSION

The pilot testing of the *WHO MNH-EPI-COS Data Collection and Analysis Toolkit* demonstrated that the Toolkit is usable, acceptable, appropriate, and feasible across diverse global research settings. Multidisciplinary teams from Brazil, Honduras, India, Rwanda, and the United Kingdom were able to implement all Toolkit components with high levels of satisfaction, drawing on retrospective datasets from four different epidemic contexts. The pilot generated a rich and actionable body of evidence that directly informed two cycles of refinement, resulting in multiple improvements to the clarity, consistency, and technical quality of all Toolkit components.

These findings support the Toolkit’s readiness for broader global dissemination and its potential to meaningfully strengthen the standardization of maternal and neonatal health data collection and reporting during epidemics. Development of multilingual versions and the prospective evaluation under real-time epidemic conditions, are the priorities that will determine whether the Toolkit achieves its ultimate goal: enabling timely, comparable, and high-quality evidence generation to protect the health of pregnant women and newborns during future epidemic threats.

## SUPPORTING INFORMATION

**S1 Table. Maternal outcomes included in datasets.**

**S2 Table. Neonatal outcomes included in datasets.**

**S3 Table. Summary of identified problems, decisions, and rationale.}**

**S1 File. Survey: team composition and dataset overview.**

**S2 File. User notes.**

**S3 File. End of pilot feedback form.**

**S4 File. Virtual consultation agenda.**

## FUNDING

This development and pilot evaluation of the *MNH-EPI-COS Data Collection and Analysis Toolkit* was conducted with funding support from the Gates Foundation (INV087293), and the UNDP/UNFPA/UNICEF/WHO/World Bank Special Programme of Research, Development and Research Training in Human Reproduction (HRP), Department of Reproductive Health and Research, World Health Organization (WHO), Geneva, Switzerland (WHO Registration 2025/1594262-0; 2026/1628205-0). The views of the funding bodies have not influenced the content of this manuscript.

## DATA AVAILABILITY

Data may be made available upon reasonable request subject to relevant data sharing agreements and ethical approvals.

## AUTHOR CONTRIBUTIONS

The *MNH-EPI-COS Data Collection and Analysis Toolki*t and its evaluation was conceptualised by VP, MBe, KK, MBel, JPA, LG and AC. VP, Mbo, KK and TD coordinated the pilot evaluation. VP and JPA conducted the analysis and drafted the first version of the manuscript. All authors (KK, TD, LG, AC, JGC, MLC, CS, RTS, JA, JGA, PB, DSM, SSG, AK, CK, MSS, LMM, PN. JCU, BU, RMS, AP, AW, MBel and MBe) reviewed and contributed substantially to the interpretation and revision of the manuscript, and approved the final version for submission.

## COMPETING INTERESTS

The authors declare no competing interests.

## ACKNOWLEDGEMENTS

We gratefully acknowledge all participating teams and experts for their valuable feedback, contributions, time, and engagement throughout the Toolkit pilot and improvements. We also sincerely thank the following individuals for their support of this project: Olufemi Oladapo and Mercedes Bonet (UNDP-UNFPA-UNICEF-WHO-World Bank Special Programme of Research, Development and Research Training in Human Reproduction, Department of Sexual and Reproductive Health and Research, World Health Organization, Geneva, Switzerland); and Ana Paula Perez Oyola, Milagros Oberti, Pablo Solares, Sandra Formia, and Carolina Nigri (Institute for Clinical Effectiveness and Health Policy, Argentina). We extend our sincere thanks to the experts who reviewed the toolkit materials. From WHO headquarters, staff contributed to content review at different stages: Vanessa Brizuela, Edna Kara, Caron Rahn Kim, Ronaldo Silva, Soe Soe Thwin (LHR); Smaragda Lamprianou (Department of Regulation and Prequalification) and Jamie Rylance (Department of the WHO Emergencies Programme).

External experts: Daniele De Luca (Université Paris-Saclay, France), Sangappa Dhaded (KLE Academy of Higher Education & Research, India), Marian Knight (University of Oxford, United Kingdom), Silke Mader (Global Foundation for the Care of Newborn Infants, Germany), Tavitiya Sudjaritruk (Chiang Mai University, Thailand), Manal Younus (Iraqi Pharmacovigilance Centre, Ministry of Health, Iraq), and Noreen Zafar (Girls and Women Health Initiative, Pakistan).

## Notes

### Competing Interest Statement

The authors have declared no competing interest.

